# The Multiple Sclerosis severity allele rs10191329^A^ and cognitive function: a UK Biobank study

**DOI:** 10.1101/2025.09.28.25336369

**Authors:** Ioanna Zimianiti, Sheena Waters, Adil Harroud, Pernilla Stridh, Ruth Dobson, Benjamin M. Jacobs

## Abstract

The genome-wide association study of Multiple Sclerosis severity linked the genetic variant rs10191329^A^ to long-term disability, suggesting it affects central nervous system resilience. We hypothesised that rs10191329^A^ might influence cognition in other neurological diseases and in healthy controls. We explored the relationship between rs10191329^A^ and cognitive function - measured using reaction time, fluid intelligence, and prospective memory tests - in the United Kingdom Biobank. Rs10191329^A^ was consistently associated with poorer cognition in controls, with similar but non-significant trends in Multiple Sclerosis, Parkinson’s disease, and dementia. Our results support the hypothesis that rs10191329^A^ affects multiple sclerosis outcomes by modulating brain health.

## Introduction

Multiple Sclerosis (MS) is an autoimmune and degenerative disease of the Central Nervous System (CNS). Preventing long-term disability accrual remains an unmet need despite effective suppression of inflammatory relapses(1). Recently, a genome-wide association study (GWAS) of >12,000 MS cases linked an intergenic variant at the *DYSF*-*ZNF638* locus, rs10191329^A^, to cross-sectional physical disability(2). Cell-type heritability enrichment implicated CNS-resident cells, and Mendelian Randomisation suggested genetic overlap with educational attainment(2). Further studies have linked rs10191329^A^ to brain atrophy, elevated serum Neurofilament light chain (NFL) levels and retinal layer atrophy, markers of neuroaxonal degeneration(3)(4)(5). Attempts to replicate the association in smaller cohorts have demonstrated consistent directions of effect(6)(7)(8). These findings reinforce a CNS-mediated mechanism of MS progression. The concept that neuronal resilience modulates disease progression has support across neurological disorders, including MS, Alzheimer’s disease (AD) and Parkinson’s Disease (PD)(9)(10)(11)(12).

We therefore hypothesised that rs10191329^A^ might influence cognition in MS, healthy controls, and other neurological disorders and tested this hypothesis using genetic and cognitive test data from UK Biobank (UKB)(13).

## Materials and methods

### Cohort

The UKB is a population-based prospective cohort study described elsewhere(13). Participants aged 40-69 were recruited between 2006-2010. Data were collected through questionnaires, interviews, physical and functional measures assessments, and biological samples(13). Participants provided informed consent and can withdraw anytime, after which their data are excluded from further analyses.

### Case-control definitions

MS status was determined using UKB “first occurrences” (Category 1712), which identify the first occurrence of a set of diagnostic codes mapped to 3-character ICD-10 codes; cases were those with the code G35. To differentiate prevalent from incident cases, accounting for diagnostic and recording lag, we evaluated whether the initial diagnostic code appeared before or within 10 years of participants’ baseline assessment(14). Similarly, we identified participants with migraine (G43), as a negative disease control. We identified individuals with PD and all-cause dementia, utilising algorithmically-defined outcomes (Category 50), which use combinations of coded information for selected conditions.

We restricted analysis to participants of European genetic ancestry (Data-Field 22006) and included prevalent cases creating five mutually-exclusive cohorts:

- MS: Participants with a MS code ≤10 years after recruitment
- PD: Participants with no code for MS and a PD code ≤10 years after recruitment
- Dementia: Participants with no code for MS or PD, but with a dementia code ≤10 years after recruitment
- Migraine: Participants with no code for MS, PD, or Dementia but with a migraine code ≤10 years after recruitment
- Controls: Participants with no code for MS, PD, dementia or migraine.

### Exposure definitions

Genotype and quality control protocols are described elsewhere(13). Genotype data for rs10191329^A^ was obtained from UKB and extracted using ‘--recode AD’ flag in PLINK version 2(15). The imputation quality score was 0.98, indicating high-quality imputation. Participants with missing values for rs10191329^A^ were excluded. The hardcall dosage for the A allele was obtained as follows:

- rs10191329^A^ allelic dosage < 0.1: non-carriers.
- rs10191329^A^ allelic dosage > 0.9 and < 1.1: heterozygous
- rs10191329^A^ allelic dosage > 1.9: homozygous.

Genetic principal components (PCs) were supplied by UKB (Data-Field 22009).

### Outcome definitions

We used self-reported disability claims (Data-Field 6146), creating a binary variable indicating whether participants received any form of disability allowance.

For our cognitive outcomes we used the tests administered at recruitment with N >100,000, excluding ‘pairs matching’ which has low test-retest reliability and correlation with formal testing(16). We assessed ‘reaction time’ (Data-Field 20023) in a computerized card-matching task, with higher scores indicating slower responses. Fluid intelligence (Data-Field 20016) was measured as the total number of correct answers out of 13, with higher scores indicating better performance. ‘Prospective memory’ (Data-Field 20018) in a task requiring participants to override a prompt and select the correct target, was assessed as a binary outcome with success defined as correct recall on first attempt. The chosen cognitive outcomes were validated by demonstrating poorer performance in MS, PD and dementia cohorts compared to controls and migraine cohorts. Details on outcome measures handling and validation are provided in Supplementary Tables 1&2 and supplementary figure 1.

### Statistical analysis

We assessed the distribution of continuous variables by inspecting histograms and applying the Kolmogorov-Smirnov test; data was described as median with interquartile range (IQR), or mean with standard deviation (SD), as appropriate.

We first validated the relationship between rs10191329^A^ and disability claims in MS using logistic regression adjusted for age, sex, and the first four PCs. We then validated our cognitive outcomes by examining their associations with prevalent neurological disease, adjusting for age and sex.

For the primary analysis, we assessed the relationship between rs10191329^A^ and cognitive outcomes using regression models stratified by disease status (separately for each cohort). We applied linear regression to reaction time and fluid intelligence, logistic regression to prospective memory, and used additive genetic models adjusted for age, sex, and the first four PCs. Linear outcomes were rank-inverse normal transformed (RINT) to meet linear-model assumptions. As this was a hypothesis-driven replication analysis we used one-tailed *P-*values.

We performed sensitivity analyses without covariate adjustment, under dominant and recessive models. We applied a false discovery rate (FDR) correction to account for multiple testing, adjusting for all terms simultaneously.

We performed power calculations, using reaction time as an example due to its large sample size. We simulated a normal distribution with mean 0 and SD 1 and performed 1,000 bootstrap simulations over different plausible effect sizes varying the number of cases. For each simulation, we used univariable linear models, regressing the genotype on reaction time. Power at the 0.05 alpha level was defined as the proportion of bootstrap iterations with two-tailed *P*-values of <0.05.

### Computing

Analysis was conducted using R version 4.2.2, and utilising Queen Mary’s Apocrita HPC(17).

## Results

### Cohort characteristics

We included 399,031 participants, comprising 373,530 controls (median age 58, IQR 12, 53% female), 2,337 dementia (median age 65, IQR 6, 47.7% female), 19,672 migraine (median age 56, IQR 13, 75.4% female), 2,026 MS (median age 56, IQR 13, 72.7% female) and 1,466 PD (median age 63, IQR 7, 39.1% female) cases. The median age at MS report was 44 (IQR 16), consistent with previous estimates(18). Demographics and participants flow are demonstrated in **supplementary table 3** and **supplementary figure 2**.

### Rs10191329^A^ correlates with disability claims in MS

We demonstrated a dose-dependent relationship between rs10191329^A^ and disability claims, validating the association between rs10191329^A^ and physical disability in MS (NMS = 2,006). Carriers of two A alleles were 5.9% more likely to self-report disability claims than common allele homozygotes. Regression analysis showed modest statistical evidence (one-tailed *P* = 0.08) (OR 1.03, 95% CI 0.99 to 1.07; **figure 1**).

**Figure 1:**
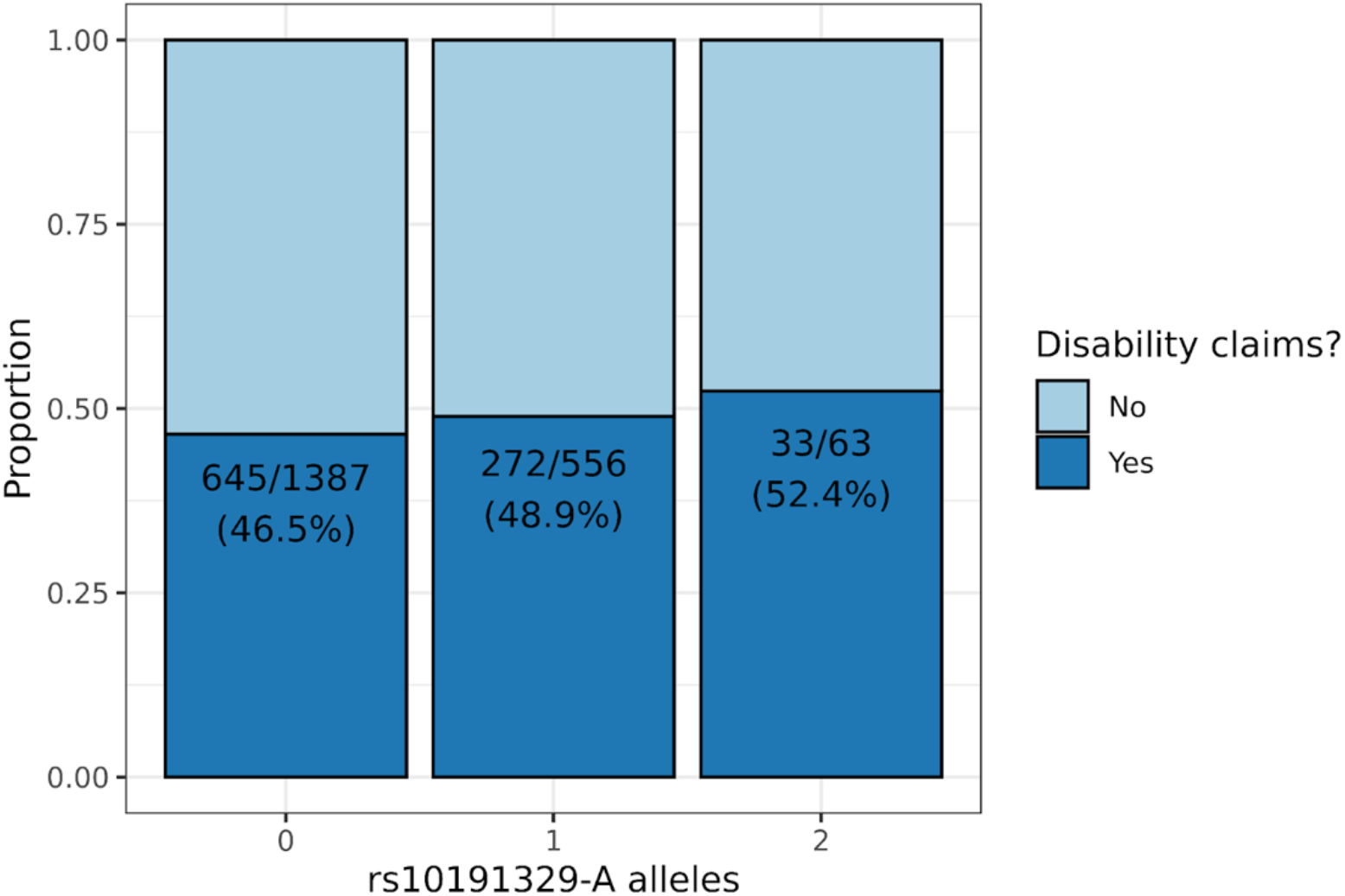
stacked barplots showing the association between rs10191329 genotype and self-reported physical disability claims among UK Biobank participants with MS. The counts shown reflect the number of individuals reporting any claim divided by the total number with MS within each genotype group. The dark blue bars indicate the proportion of individuals making at least one type of disability claim, and the light blue indicate the proportion making no claims.

### Rs10191329^A^ correlates with impaired cognition

Rs10191329^A^ was associated with slower reaction time (***β***= 0.01, 95% CI 0.003 to 0.02, *P* = 0.001), lower fluid intelligence (***β***= -0.02, 95% CI -0.03 to -0.01, *P* = 0.0004), and higher error rates on the prospective memory task (OR 1.06, 95% CI 1.03 to 1.09, *P* = 0.00002) in controls (**figure 2**). No other associations reached study-wide significance (FDR < 5%). However, we observed directionally-concordant associations between rs10191329^A^ and slower reaction time in MS, PD, and dementia, with lower fluid intelligence in all cohorts, and impaired prospective memory in dementia. Results were consistent after sensitivity analysis **(Supplementary table 4)**.

**Figure 2:**
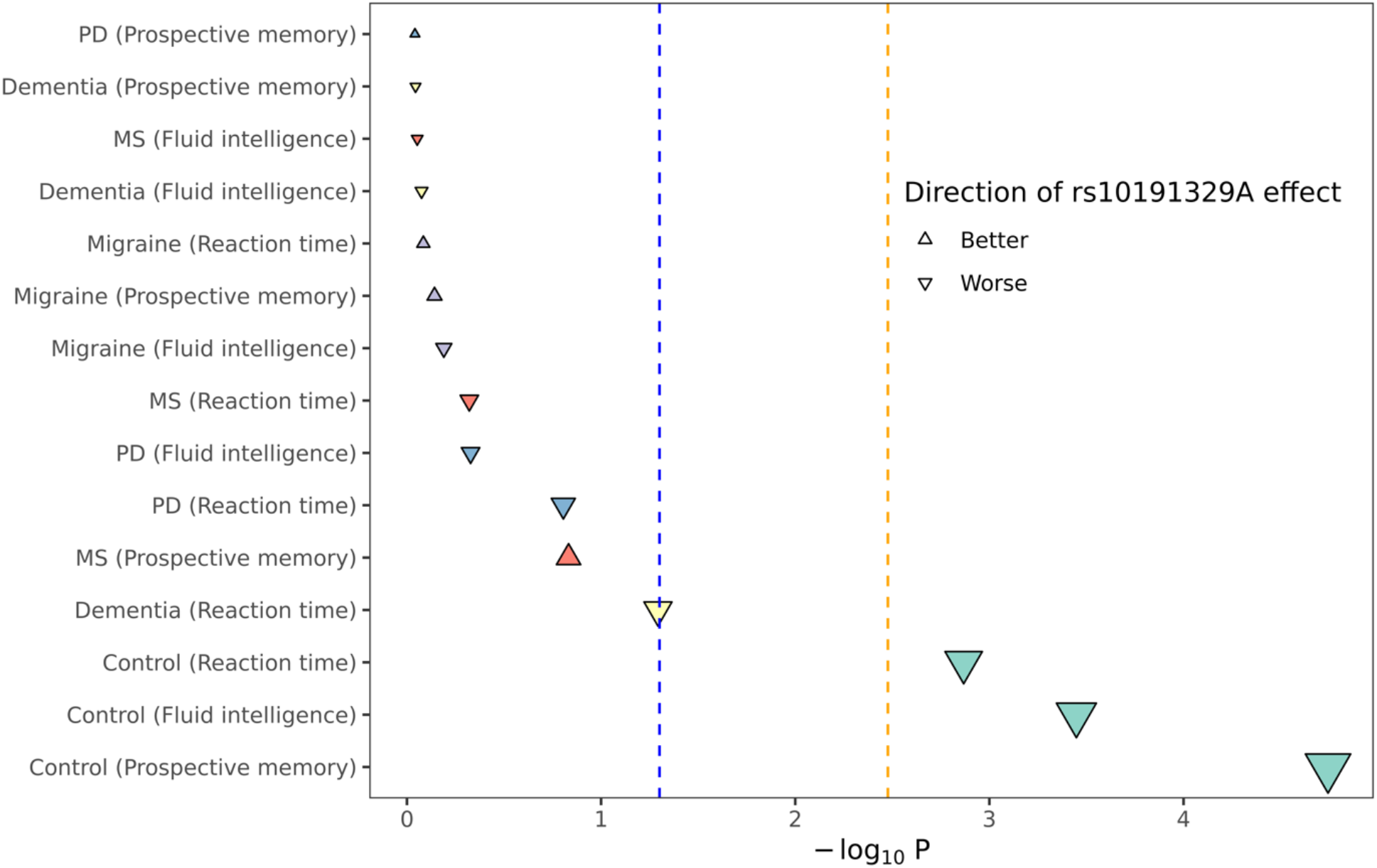
plot showing the effect directions and -log10(P) values from regression models assessing the relationship between rs10191329 genotype and cognitive outcomes in each disease group. Each point represents a regression coefficient. The shape of the point indicates the effect direction, with downward triangles indicating that rs10193219^A^ was associated with worse cognition, and vice-versa.. The colour of the dots indicates the disease group, and the y axis indicates the specific coefficient (i.e. combination of disease group and test). Points are ordered by their strength of association such that the strongest signals (i.e. largest -log10(P) values) are at the bottom of the plot. The x axis shows the strength of association, as does the size of the point. The dotted blue and orange vertical lines indicate P = 0.05 and P_Bonferroni_ = 0.05 respectively. Model coefficients are adjusted for age, sex and the first four PCs.

### Power calculations

Given that the observational difference in reaction time between dementia cases and controls is ∼0.5 SD, a per-allele increase of 0.25 SD is an upper ceiling for a plausible effect (Dementia: mean 635ms, SD 162ms; Controls: mean 555ms, SD 112ms). Power calculations indicated we can detect a per-allele increase in reaction time of >0.15 SD (97% power) in the MS cohort (n = 1,993), but not weaker associations (e.g. 66% power for a 0.1-SD per-allele increase). We can therefore only exclude an extreme effect of a magnitude equivalent to having dementia.

## Discussion

Rs10191329^A^ - a modifier of physical disability in MS - is associated with impaired cognitive outcomes in ∼370,000 healthy adults. Despite limited power to detect analogous effects in MS or other neurological disorders, we demonstrated directionally-concordant associations with cognition across health and disease.

Prior evidence has implicated rs10191329^A^ in the progression of MS, correlating the minor (A) allele with greater physical disability, radiological disease burden, neuropathological disease burden at post-mortem, and serological evidence – neurofilament light chain – of increased neurodegeneration in MS(2)(3)(4)(19). Our results – demonstrating association between rs10191329^A^ and impaired cognitive performance in a large cohort of healthy UK adults - indicate that such effects may extend beyond MS, arguing that rs10191329^A^ may influence cognition across health and disease through shared mechanisms(2)(4).

Limitations of this study include a limited cognitive test set, absence of disease-specific outcomes, limited power, and lack of a replication dataset. To minimise selection bias we focussed on tests administered to a large number of participants, but we do not explore longitudinal outcomes.

In summary, we provide evidence from population-scale cognitive testing of >370,000 adults that rs10191329^A^ is associated with lower cognitive outcomes in the general population, supporting the hypothesis that this variant influences MS severity by modulating brain health. Further work in large disease-specific cohorts is required to explore the impact of rs10191329^A^ on cognition in MS, other neurological disorders, and in health.

## Data Availability

All code for analysis is provided on: https://github.com/izimianiti/UKB-MS-severity-allele-cognition. UK Biobank data are available on request from https://www.ukbiobank.ac.uk/.

https://www.ukbiobank.ac.uk/.

## Data and code availability

All code for analysis is provided on: https://github.com/izimianiti/UKB-MS-severity-allele-cognition. UK Biobank data are available on request from https://www.ukbiobank.ac.uk/. This research was conducted under approved application 78867.

## Author contributions

All authors contributed to the manuscript. IZ, BMJ and RD contributed to the conception and design of the study. IZ, SW, BMJ and RD contributed to the acquisition and analysis of data. IZ, BMJ and RD contributed to drafting the text or preparing the figures. All authors contributed to revising the manuscript for submission.

