## Supplementary material for "The Multiple Sclerosis severity allele rs10191329^A^ and cognitive function: a UK Biobank study"

| **Supplementary table 1:** Description of outcome measures used | | |
| --- | --- | --- |
| **Outcome** | **UK Biobank Data-Fields** | **Description** |
| Disability claims (attendance/  disability/  Mobility allowance) | 6146 | Disability allowance was ascertained using the touchscreen question “Do you receive any of the following?”. Participants could select one or more of the following answers: “Attendance allowance”, “Disability living allowance”, “Blue badge”, “None of the above”, “Do not know” and “Prefer not to answer”.  We used responses at study recruitment to create a binary variable indicating whether participants received any form of disability allowance. Responses ‘prefer not to answer’ or ‘do not know’ were treated as missing. |
| Reaction Time | 20023 | Participants were shown two cards at a time over 12 rounds. We used the mean time to correctly identify matches taken from rounds 5-12 (rounds 1-4 were training). Times below 50ms were excluded as they are presumed to result from anticipation rather than reaction. Times above 2000ms were curtailed to 2000ms as the cards had disappeared by this point. A higher score is worse, reflecting slower reaction time. |
| Fluid intelligence score | 20016 | Fluid intelligence, the ability to address problems requiring logic and reasoning without the use of prior knowledge, was assessed using 13 multiple choice questions on the touch-screen. Participants were given 2 minutes to answer as many questions as possible. Each of the multiple choice questions assessed verbal (e.g. “Bud is to flower as child is to?”, Possible answers: “Grow”, “Develop”, “Improve”, “Adult”, “Old”, “Do not know”, “Prefer not to answer”) and numerical (e.g. “Add the following numbers together: 1 2 3 4 5 – is the answer?”, Possible answers: “13” “14”, “15”, “16”, “17”, “Do not know”, “Prefer not to answer”) skills. Each question had 5 possible answers that participants could choose from, in addition to options “Do not know” and “Prefer not to answer”.  Data-field  20016 represents the total number of correct answers out of 13 questions, with unattempted questions scored as 0.   Higher scores indicate better performance |
| Prospective memory score | 20018 | Participants were shown the following message on the touchscreen questionnaire before completing any of the following tests: “At the end of the games we will show you four coloured shapes and ask you to touch the Blue Square. However, to test your memory, we want you to actually touch the Orange Circle instead”. After the reaction time test participants were shown a series of shapes and were asked to touch the blue square. If participants touched the blue square the following prompt appeared: “At the start of the games we asked you to remember to touch a different symbol when this screen appeared. Please try to remember which symbol it was and touch it now” Data field 20018 represents whether participants answered the question correctly on the first attempt, on the second attempt or whether the instruction was not recalled, was skipped or was incorrect. We assessed prospective memory score as a binary outcome, considering recall on first attempt as a positive outcome, and other outcomes (correct recall on second attempt, instruction forgotten or incorrect response) as a failure. |

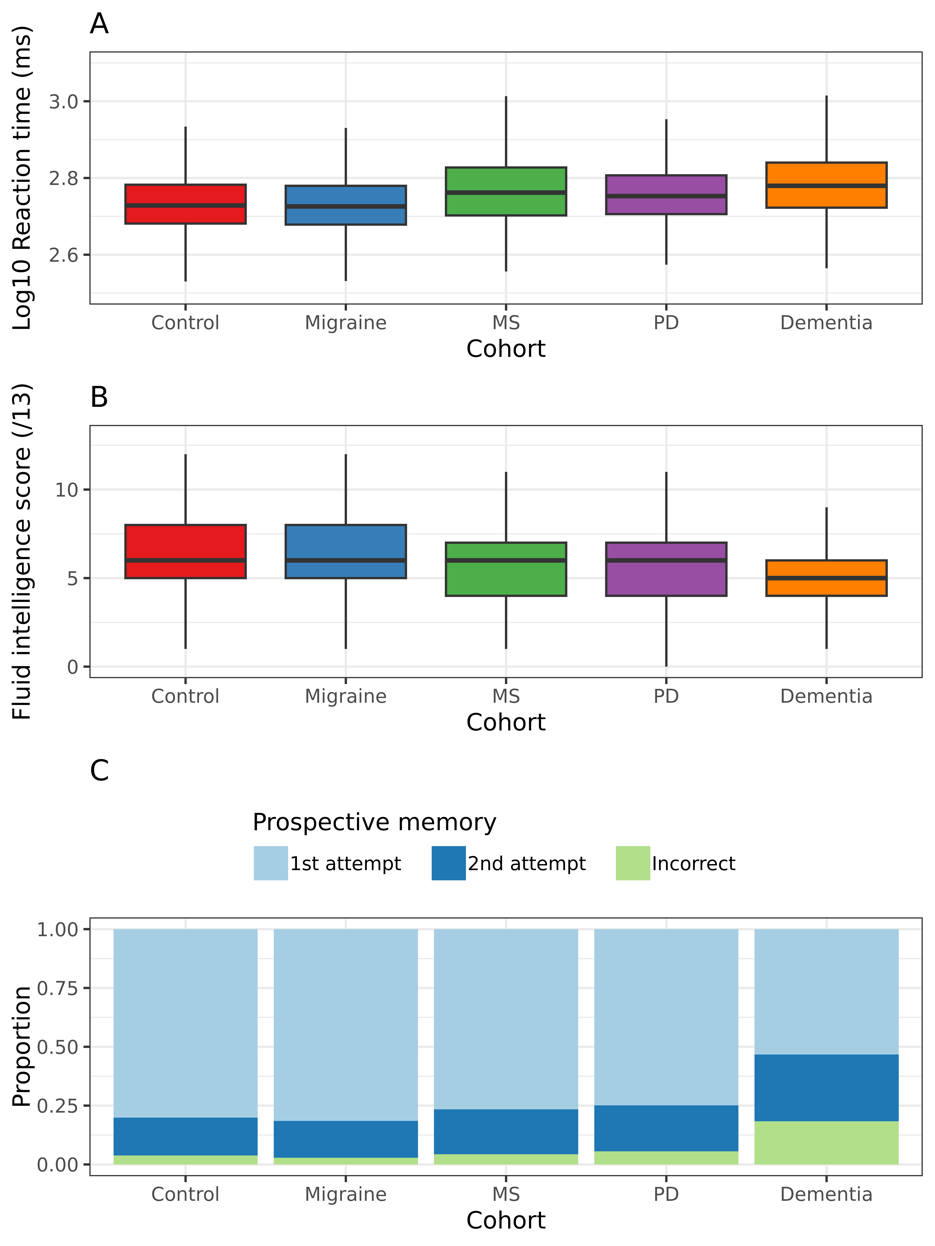

***Supplementary Figure 1*** *- boxplots displaying the reaction time (in ms) for UK Biobank participants in each of the mutually-excluded disease cohorts (x axis), For clarity the y axis shows reaction times on the log10 scale. Higher reaction times indicate poorer performance. B - as per A, but showing the raw fluid intelligence scores for each cohort. Scores are shown out of 13, with higher scores indicating better performance. C - stacked barplots showing the performance of each disease cohort on a prospective memory task. The outcomes shown are incorrect (the worst outcome), correct on the 1st attempt (best outcome), and correct on the 2nd attempt. A higher proportion of respondents correct at the 1st attempt indicates better performance.*

| Supplementary Table 2: associations between prevalent neurological diseases and cognitive test scores. The table shows the output of multivariable linear regression (for reaction time and fluid intelligence) or logistic regression (for prospective memory) models adjusted for age and sex. The beta coefficients displayed reflect the association between the stated disease cohort and the cognitive test score, with healthy controls as the reference category. For each outcome, the direction of effect is simplified as 'better' or 'worse' than controls. For reaction time and fluid intelligence, the outcome was subjected to rank inverse normalisation prior to model fitting and so the beta coefficient is on this scale. For prospective memory, an incorrect response was coded as a '1' and a correct response as a '0', and so the beta coefficient refers to the log(Odds Ratio) of an incorrect response, i.e. beta > 0 ~ worse performance. False Discovery Rate (FDR) adjusted P values reflect a global FDR, accounting for a total of 21 terms (intercept, age, sex, and 4 disease terms). FDR values with < 0.05 are indicated with a *. | | | | | | |
| --- | --- | --- | --- | --- | --- | --- |
| **Cohort** | **Direction** | **Beta** | **SE** | **P** | **FDR** | **Significance** |
| **Reaction time** | | | | | | |
| Migraine | Worse | 0.000 | 0.007 | 0.958 | 0.958 |  |
| MS | Worse | 0.466 | 0.021 | 1.22E-108 | 1.46E-107 | * |
| PD | Worse | 0.132 | 0.025 | 9.82E-08 | 2.36E-07 | * |
| Dementia | Worse | 0.337 | 0.020 | 2.11E-64 | 1.27E-63 | * |
| **Fluid intelligence** | | | | | | |
| Migraine | Better | 0.041 | 0.013 | 0.002 | 0.004 | * |
| MS | Worse | -0.131 | 0.039 | 8.91E-04 | 0.002 | * |
| PD | Worse | -0.055 | 0.045 | 0.226 | 0.272 |  |
| Dementia | Worse | -0.501 | 0.037 | 1.13E-42 | 3.39E-42 | * |
| **Prospective memory** | | | | | | |
| Migraine | Better | -0.048 | 0.035 | 0.172 | 0.229 |  |
| MS | Worse | 0.253 | 0.094 | 0.007 | 0.011 | * |
| PD | Worse | 0.112 | 0.106 | 0.287 | 0.314 |  |
| Dementia | Worse | 0.999 | 0.072 | 3.00E-44 | 1.20E-43 | * |

| Supplementary Table 3: demographics and raw cognitive test scores of included participants, stratified by disease cohort. Quantitative variables are presented as median and interquartile range, categorical variables are presented as n and %. | | | | | |
| --- | --- | --- | --- | --- | --- |
| **Variable** | **Control** | **Migraine** | **MS** | **PD** | **Dementia** |
| **Total N** | 373,530 | 19,672 | 2,026 | 1,466 | 2,337 |
| **MAF** | 17.5% | 17.5% | 16.9% | 17.2% | 17.5% |
| **N for each cognitive test** |  |  |  |  |  |
| Reaction time | 371,364 | 19,583 | 1,993 | 1,447 | 2,245 |
| Fluid intelligence | 120,425 | 5,618 | 615 | 467 | 717 |
| Prospective memory | 122,816 | 5,701 | 645 | 485 | 807 |
| **Age at baseline** | 58 (12) | 56 (13) | 56 (13) | 63 (7) | 65 (6) |
| **Age at diagnosis** | N/A | 38(30) | 44(16) | 66(12) | 72(8) |
| **Gender** |  |  |  |  |  |
| Female | 197,857 (53%) | 14,838 (75.4%) | 1,473 (72.7%) | 573 (39.1%) | 1,115 (47.7%) |
| Male | 175,673 (47%) | 4,834 (24.6%) | 553 (27.3%) | 893 (60.9%) | 1,222 (52.3%) |
| **rs10191329 genotype** |  |  |  |  |  |
| C/C | 254,130 (68%) | 13,390 (68.1%) | 1,404 (69.3%) | 1,006 (68.6%) | 1,583 (67.7%) |
| C/A | 107,847 (28.9%) | 5,665 (28.8%) | 558 (27.5%) | 415 (28.3%) | 689 (29.5%) |
| A/A | 11,553 (3.1%) | 617 (3.1%) | 64 (3.2%) | 45 (3.1%) | 65 (2.8%) |
| **Prospective memory** |  |  |  |  |  |
| Not recalled/skipped/incorrect | 4,705 (3.8%) | 162 (2.8%) | 28 (4.3%) | 27 (5.6%) | 148 (18.3%) |
| Correct recall on first attempt | 98,378 (80.1%) | 4,648 (81.5%) | 494 (76.6%) | 363 (74.8%) | 430 (53.3%) |
| Correct recall on second attempt | 1,9733 (16.1%) | 891 (15.6%) | 123 (19.1%) | 95 (19.6%) | 229 (28.4%) |
| **Reaction time (ms)** | 535 (126) | 532 (125) | 578 (172) | 566 (133) | 602 (168) |
| **Fluid intelligence score (/13)** | 6 (3) | 6 (3) | 6 (3) | 6 (3) | 5 (2) |

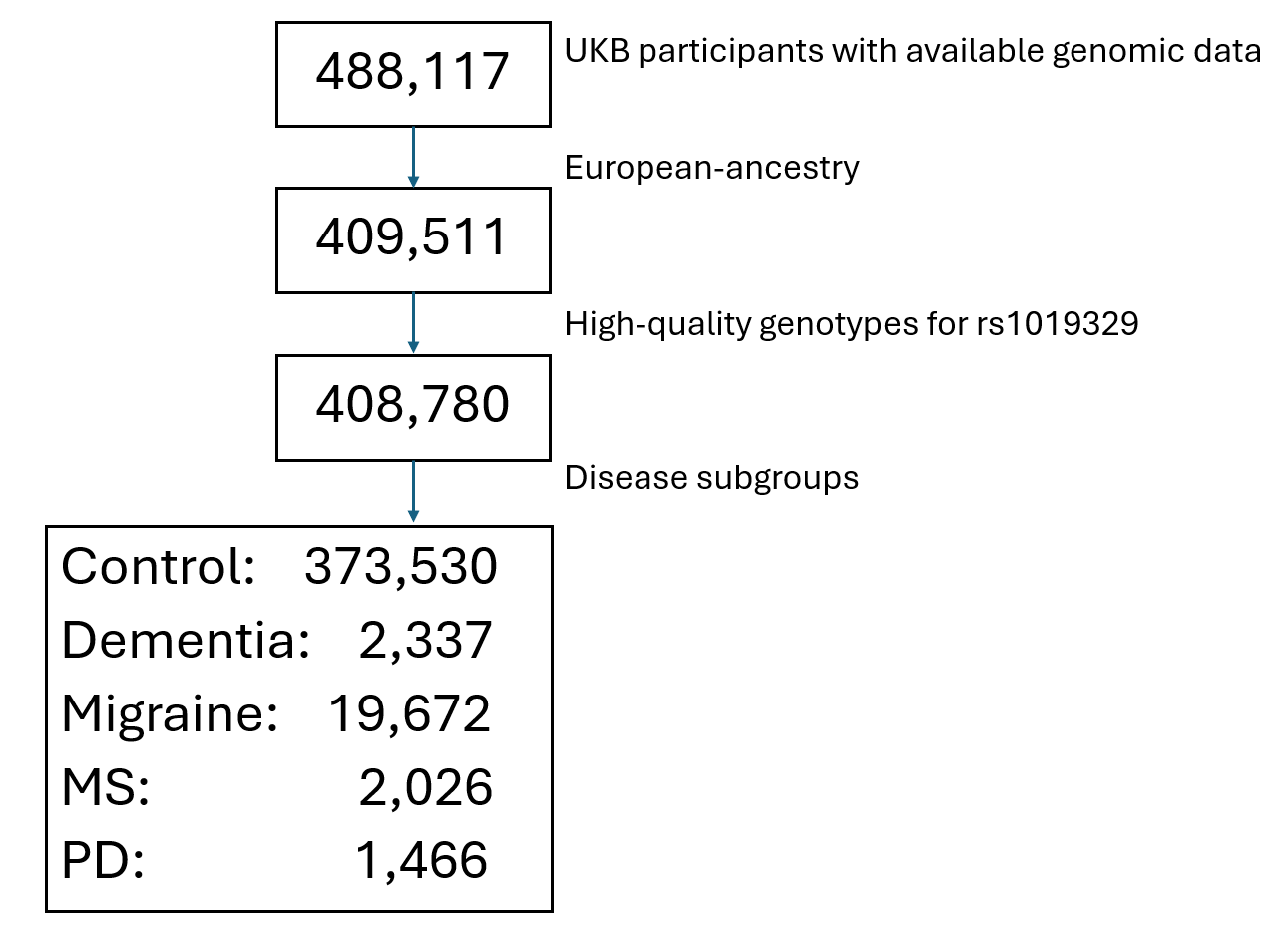

***Supplementary Figure 2:*** *flow diagram indicating the numbers of participants at each stage of the inclusion and exclusion criteria. In total the final analysis cohort comprised 399,031 White British UKB participants.*

| Supplementary table 4: the table shows the association between rs10191329 genotype and cognitive test performance in each disease cohort. Models were adjusted for age, sex, and the first four genetic principal components. The 'genotype N' column shows the number within each disease cohort for the stated test with each of the three genotypes (CC, CA, or AA). Sensitivity analyses were performed including an unadjusted model, use of a dominant genetic model, and use of a recessive model. | | | | | | | | | | | |
| --- | --- | --- | --- | --- | --- | --- | --- | --- | --- | --- | --- |
| **Cohort** | **Cognitive test** | **Genotype N (CC/CA/AA)** | **Model** | **Beta** | **SE** | **Lower CI** | **Upper CI** | **P** | **FDR** | **Direction** | **Significance** |
| Control | Reaction time | N = 252634/107251/11479 | Adjusted | 0.009 | 0.003 | 0.004 | 0.015 | 0.001 | 0.010 | Worse | * |
| Migraine | Reaction time | N = 13324/5643/616 | Adjusted | -0.003 | 0.012 | -0.027 | 0.021 | 0.823 | 0.958 | Better |  |
| MS | Reaction time | N = 1381/549/63 | Adjusted | 0.033 | 0.046 | -0.057 | 0.123 | 0.477 | 0.875 | Worse |  |
| Dementia | Reaction time | N = 1519/661/65 | Adjusted | 0.079 | 0.040 | 0.000 | 0.157 | 0.051 | 0.218 | Worse |  |
| PD | Reaction time | N = 993/410/44 | Adjusted | 0.067 | 0.047 | -0.026 | 0.160 | 0.157 | 0.494 | Worse |  |
| Control | Fluid intelligence | N = 82026/34673/3726 | Adjusted | -0.019 | 0.005 | -0.029 | -0.008 | 0.000 | 0.004 | Worse | * |
| MS | Fluid intelligence | N = 431/162/22 | Adjusted | -0.010 | 0.068 | -0.142 | 0.123 | 0.885 | 0.958 | Worse |  |
| Migraine | Fluid intelligence | N = 3864/1566/188 | Adjusted | -0.011 | 0.024 | -0.058 | 0.036 | 0.646 | 0.953 | Worse |  |
| PD | Fluid intelligence | N = 316/135/16 | Adjusted | -0.064 | 0.089 | -0.238 | 0.110 | 0.470 | 0.875 | Worse |  |
| Dementia | Fluid intelligence | N = 501/201/15 | Adjusted | -0.014 | 0.071 | -0.154 | 0.126 | 0.841 | 0.958 | Worse |  |
| Control | Prospective memory | N = 83627/35382/3807 | Adjusted | 0.057 | 0.013 | 0.031 | 0.083 | 0.000 | 0.001 | Worse | * |
| MS | Prospective memory | N = 455/167/23 | Adjusted | -0.269 | 0.185 | -0.632 | 0.095 | 0.147 | 0.490 | Better |  |
| Migraine | Prospective memory | N = 3926/1586/189 | Adjusted | -0.023 | 0.064 | -0.148 | 0.102 | 0.721 | 0.958 | Better |  |
| PD | Prospective memory | N = 328/141/16 | Adjusted | -0.023 | 0.198 | -0.411 | 0.366 | 0.909 | 0.958 | Better |  |
| Dementia | Prospective memory | N = 564/227/16 | Adjusted | 0.017 | 0.141 | -0.259 | 0.293 | 0.903 | 0.958 | Worse |  |
| Control | Reaction time | N = 252634/107251/11479 | Dominant | 0.010 | 0.003 | 0.004 | 0.017 | 0.001 | 0.010 | Worse | * |
| Migraine | Reaction time | N = 13324/5643/616 | Dominant | -0.001 | 0.014 | -0.029 | 0.026 | 0.933 | 0.958 | Better |  |
| MS | Reaction time | N = 1381/549/63 | Dominant | 0.040 | 0.053 | -0.065 | 0.145 | 0.453 | 0.875 | Worse |  |
| Dementia | Reaction time | N = 1519/661/65 | Dominant | 0.091 | 0.046 | 0.001 | 0.181 | 0.048 | 0.218 | Worse |  |
| PD | Reaction time | N = 993/410/44 | Dominant | 0.116 | 0.055 | 0.009 | 0.223 | 0.033 | 0.170 | Worse |  |
| Control | Fluid intelligence | N = 82026/34673/3726 | Dominant | -0.022 | 0.006 | -0.034 | -0.010 | 0.000 | 0.004 | Worse | * |
| MS | Fluid intelligence | N = 431/162/22 | Dominant | 0.009 | 0.080 | -0.148 | 0.165 | 0.913 | 0.958 | Better |  |
| Migraine | Fluid intelligence | N = 3864/1566/188 | Dominant | -0.008 | 0.028 | -0.063 | 0.047 | 0.772 | 0.958 | Worse |  |
| PD | Fluid intelligence | N = 316/135/16 | Dominant | -0.035 | 0.104 | -0.237 | 0.168 | 0.739 | 0.958 | Worse |  |
| Dementia | Fluid intelligence | N = 501/201/15 | Dominant | -0.043 | 0.080 | -0.199 | 0.113 | 0.591 | 0.953 | Worse |  |
| Control | Prospective memory | N = 83627/35382/3807 | Dominant | 0.054 | 0.015 | 0.024 | 0.084 | 0.000 | 0.004 | Worse | * |
| MS | Prospective memory | N = 455/167/23 | Dominant | -0.264 | 0.214 | -0.683 | 0.156 | 0.218 | 0.526 | Better |  |
| Migraine | Prospective memory | N = 3926/1586/189 | Dominant | 0.005 | 0.074 | -0.141 | 0.150 | 0.951 | 0.958 | Worse |  |
| PD | Prospective memory | N = 328/141/16 | Dominant | -0.157 | 0.230 | -0.609 | 0.295 | 0.496 | 0.875 | Better |  |
| Dementia | Prospective memory | N = 564/227/16 | Dominant | -0.065 | 0.156 | -0.371 | 0.240 | 0.675 | 0.958 | Better |  |
| Control | Reaction time | N = 252634/107251/11479 | Recessive | 0.012 | 0.009 | -0.005 | 0.030 | 0.166 | 0.498 | Worse |  |
| Migraine | Reaction time | N = 13324/5643/616 | Recessive | -0.018 | 0.038 | -0.092 | 0.056 | 0.641 | 0.953 | Better |  |
| MS | Reaction time | N = 1381/549/63 | Recessive | 0.028 | 0.141 | -0.248 | 0.304 | 0.844 | 0.958 | Worse |  |
| Dementia | Reaction time | N = 1519/661/65 | Recessive | 0.092 | 0.128 | -0.160 | 0.344 | 0.473 | 0.875 | Worse |  |
| PD | Reaction time | N = 993/410/44 | Recessive | -0.198 | 0.148 | -0.488 | 0.093 | 0.182 | 0.512 | Better |  |
| Control | Fluid intelligence | N = 82026/34673/3726 | Recessive | -0.020 | 0.016 | -0.052 | 0.012 | 0.219 | 0.526 | Worse |  |
| MS | Fluid intelligence | N = 431/162/22 | Recessive | -0.138 | 0.198 | -0.526 | 0.250 | 0.487 | 0.875 | Worse |  |
| Migraine | Fluid intelligence | N = 3864/1566/188 | Recessive | -0.046 | 0.072 | -0.188 | 0.095 | 0.523 | 0.897 | Worse |  |
| PD | Fluid intelligence | N = 316/135/16 | Recessive | -0.352 | 0.267 | -0.876 | 0.171 | 0.188 | 0.512 | Worse |  |
| Dementia | Fluid intelligence | N = 501/201/15 | Recessive | 0.256 | 0.255 | -0.243 | 0.755 | 0.315 | 0.676 | Better |  |
| Control | Prospective memory | N = 83627/35382/3807 | Recessive | 0.157 | 0.040 | 0.079 | 0.235 | 0.000 | 0.002 | Worse | * |
| MS | Prospective memory | N = 455/167/23 | Recessive | -0.752 | 0.634 | -1.996 | 0.491 | 0.236 | 0.544 | Better |  |
| Migraine | Prospective memory | N = 3926/1586/189 | Recessive | -0.253 | 0.206 | -0.657 | 0.151 | 0.219 | 0.526 | Better |  |
| PD | Prospective memory | N = 328/141/16 | Recessive | 0.798 | 0.543 | -0.266 | 1.862 | 0.141 | 0.490 | Worse |  |
| Dementia | Prospective memory | N = 564/227/16 | Recessive | 0.959 | 0.548 | -0.115 | 2.032 | 0.080 | 0.320 | Worse |  |
| Control | Reaction time | N = 252634/107251/11479 | Unadjusted | 0.009 | 0.003 | 0.003 | 0.015 | 0.002 | 0.014 | Worse | * |
| Migraine | Reaction time | N = 13324/5643/616 | Unadjusted | 0.002 | 0.013 | -0.023 | 0.027 | 0.873 | 0.958 | Worse |  |
| MS | Reaction time | N = 1381/549/63 | Unadjusted | 0.028 | 0.047 | -0.065 | 0.121 | 0.549 | 0.915 | Worse |  |
| Dementia | Reaction time | N = 1519/661/65 | Unadjusted | 0.087 | 0.041 | 0.007 | 0.167 | 0.034 | 0.170 | Worse |  |
| PD | Reaction time | N = 993/410/44 | Unadjusted | 0.050 | 0.048 | -0.045 | 0.144 | 0.304 | 0.676 | Worse |  |
| Control | Fluid intelligence | N = 82026/34673/3726 | Unadjusted | -0.018 | 0.005 | -0.029 | -0.008 | 0.000 | 0.004 | Worse | * |
| MS | Fluid intelligence | N = 431/162/22 | Unadjusted | -0.004 | 0.067 | -0.136 | 0.129 | 0.958 | 0.958 | Worse |  |
| Migraine | Fluid intelligence | N = 3864/1566/188 | Unadjusted | -0.011 | 0.024 | -0.058 | 0.036 | 0.651 | 0.953 | Worse |  |
| PD | Fluid intelligence | N = 316/135/16 | Unadjusted | -0.045 | 0.089 | -0.219 | 0.128 | 0.608 | 0.953 | Worse |  |
| Dementia | Fluid intelligence | N = 501/201/15 | Unadjusted | -0.022 | 0.071 | -0.162 | 0.118 | 0.760 | 0.958 | Worse |  |
| Control | Prospective memory | N = 83627/35382/3807 | Unadjusted | 0.054 | 0.013 | 0.028 | 0.080 | 0.000 | 0.001 | Worse | * |
| MS | Prospective memory | N = 455/167/23 | Unadjusted | -0.278 | 0.183 | -0.638 | 0.081 | 0.129 | 0.483 | Better |  |
| Migraine | Prospective memory | N = 3926/1586/189 | Unadjusted | -0.019 | 0.063 | -0.143 | 0.105 | 0.764 | 0.958 | Better |  |
| PD | Prospective memory | N = 328/141/16 | Unadjusted | -0.057 | 0.194 | -0.438 | 0.324 | 0.770 | 0.958 | Better |  |
| Dementia | Prospective memory | N = 564/227/16 | Unadjusted | 0.019 | 0.139 | -0.253 | 0.292 | 0.889 | 0.958 | Worse |  |
